# Benchmarking non-additive genetic effects on polygenic prediction and machine learning-based approaches

**DOI:** 10.1101/2025.10.10.25337750

**Authors:** Nathaniel Yates Bell, Douglas P Wightman, Christiaan de Leeuw, Danielle Posthuma

**Affiliations:** Department of Complex Trait Genetics, Center for Neurogenomics and Cognitive Research, Amsterdam Neuroscience, Vrije Universiteit Amsterdam, 1081 HV, Amsterdam, The Netherlands; Department of Child and Adolescent Psychiatry and Pediatric Psychology, Section Complex Trait Genetics, Amsterdam Neuroscience, Vrije Universiteit Medical Center, 1081 HV, Amsterdam, The Netherlands

**Keywords:** genome-wide association study, dominance, polygenic scores, deep learning, machine learning

## Abstract

Polygenic scores (PGSs) are widely used to translate genome-wide association study (GWAS) findings into tools for genetic risk prediction. Most current approaches assume additive effects, yet the contribution of non-additive variation to predictive performance remains unclear. Here we investigate the impact of dominance deviations on polygenic prediction using simulated phenotypes and ten complex traits from the UK Biobank. We compare four approaches: a standard additive PGS, a dominance-adjusted PGS, gradient-boosted decision trees, and neural networks. Across most scenarios, the additive PGS performed robustly, but its accuracy declined when traits had low polygenicity, high SNP heritability, and substantial dominance effects. These results delineate the conditions under which additive models suffice and highlight when more flexible machine learning methods may be advantageous.

## Introduction

Genome-wide association studies (GWAS) have identified countless genetic variants associated with complex traits and diseases. Although translating these discoveries into biological insight or improved clinical care remains challenging^1–3^, identified variants can already be leveraged to optimize disease risk prediction by using polygenic scores (PGS). A PGS aggregates the effects of many variants into a single weighted score representing an individual’s genetic predisposition^4–6^. It is typically computed by summing the risk allele count for each disease-associated single nucleotide polymorphism (SNP), and weighting each by its effect size estimated from GWAS. The ability to predict individual disease-risk prior to diagnosis allows target groups of individuals to be identified for early screening and prevention programs, possibly minimizing the impact of disease or preventing disease occurrence^7^. Indeed, previous research has shown that stratifying individuals by PGS can identify individuals whose polygenic risk for phenotypes like coronary artery disease and type 2 diabetes is comparable to individuals with high effect mutations for monogenic disorders^8^. Therefore, PGS scores are becoming increasingly used in clinical care, for example, in risk tools and pilot programs for cancer, cardiovascular disease, and other conditions^9^. Consequently, improving the predictive accuracy of PGS is important and has been a major focus of research for the past decade.

Most state-of-the-art PGS methods (e.g., LDPred, LDPred2, Lassosum, PRS-CS, and SBayesR) all assume additive genetic risk, where each allele’s contribution is assumed to be independent from the contribution of other alleles, and to increase linearly^10–15^. This additive framework has been proven to be extremely robust in human data, with empirical studies demonstrating that additive genetic variance explains the vast majority of SNP heritability (*h*^2^). Moreover, non-additive genetic effects are thought to be small, with a recent analysis of over 1,000 phenotypes in the UK Biobank finding that on average dominant genetic effects only contribute 0.83% of the additive heritability, with prior twin studies finding this value closer to 0.5%^16–18^. However, while the average contribution of dominance to phenotypic variance is small, certain traits may have high levels of dominance and adequately modelling these effects may significantly enhance PGS prediction for those traits. Similarly, higher-order gene-gene interactions (epistasis) are well documented in model organisms and seem to contribute to human traits (particularly those under strong natural selection), however, detecting and modelling epistasis in humans is difficult due to limited power and confounding LD^19,20^.

Recent years have seen growing interest in machine learning (ML) and deep learning (DL) approaches for polygenic prediction, as these approaches are well-suited to model interdependent and non-linear genetic effects in high dimensional data^21–24^. Several studies have applied ML/DL models to construct polygenic predictors with the goal of capturing both additive and non-additive genetic signals, and some show encouraging improvements: a neural network-based PGS for breast cancer achieved higher accuracy than traditional PGS and other ML models, and a polygenic neural network model for predicting Alzheimer’s disease risk outperformed both conventional PGS and graph-based methods^21,23^. However, overall the performance gains from ML/DL-based PGS methods have been inconsistent and modest; some studies report improvements with ML/DL approaches, and others find superior predictive accuracy with additive PGS methods^25–29^.

Recent work has robustly investigated when non-additivity from epistasis (i.e., SNP-SNP interactions) may warrant the use of DL over linear PGS methods, and concluded that the utility of DL for polygenic prediction is currently limited under these settings, with any true epistatic effects likely to be small and confounded by LD-induced “joint tagging” artifacts^24^. However, it is still unknown how the impact of non-additivity at the individual SNP level – such as from dominance deviations, where the heterozygote effect on the phenotype deviates from the midpoint between the homozygotes – can impact additive PGS performance, with only one study attempting to explicitly account for these forms of non-additivity^30^. In sum, while the additive polygenic model has proven highly effective for complex traits, it remains unclear whether – and under which conditions – explicitly modelling non-additive SNP effects improves disease risk prediction.

In this study, we expand on the previous work in this area by specifically examining how non-additive SNP effects due to dominance deviations can impact additive PGS methods, and if ML/DL models may be preferrable under these conditions^24^. We address this question by simulating genotype-phenotype datasets with controlled degrees of SNP *h*^2^ attributable to causal SNPs with dominance deviations (from here referred to as “dominance-SNPs”) and comparing the predictive performance of two ML/DL models – XGBoost and a deep neural network – to additive and dominance-adjusted PGS methods in these various scenarios. Additionally, we test the models in the UK Biobank using phenotypes with known dominance effects. Through these analyses, we aim to delineate the thresholds at which heritability due to dominance-SNPs begins to have meaningful impact on additive PGS methods and examine whether ML/DL models can improve genetic prediction by capturing non-additive effects.

## Results

### Methods overview

We simulated two genotype samples with HAPNEST using the 1000G European reference panel: a discovery sample (*n* = 100,000) and a target sample (*n* = 50,000; Methods). For each data set, we simulated a range of phenotypes that varied in terms of (i) total SNP *h*^2^ (10%, 20, 50%); (ii) proportion of the SNP *h*^2^ attributable to causal dominance-SNPs (0% - 100%); (iii) the number of causal SNPs (100 – 1000); and (iv), the magnitude of the dominance deviation (*k*), with a range of 0 to -1.0 (Methods; Table S1 – S2). We compared four models: (a) a linear regression with an additive PGS score (ADD PGS); (b) a linear regression using a dominance-adjusted PGS score (DOM PGS); (c) a deep neural network (DNN); and (d) an XGBoost ensemble learning model (XGB; Figure 1). Models were run using fully nested 5-fold cross validation, and the mean variance-explained (*R*^2^) across the 5 test folds was used to evaluate their performance (Fig. 1). Lastly, we applied the 4 models to 10 traits in the UK Biobank to examine the effects of dominance deviations in real data.

**Figure 1:**
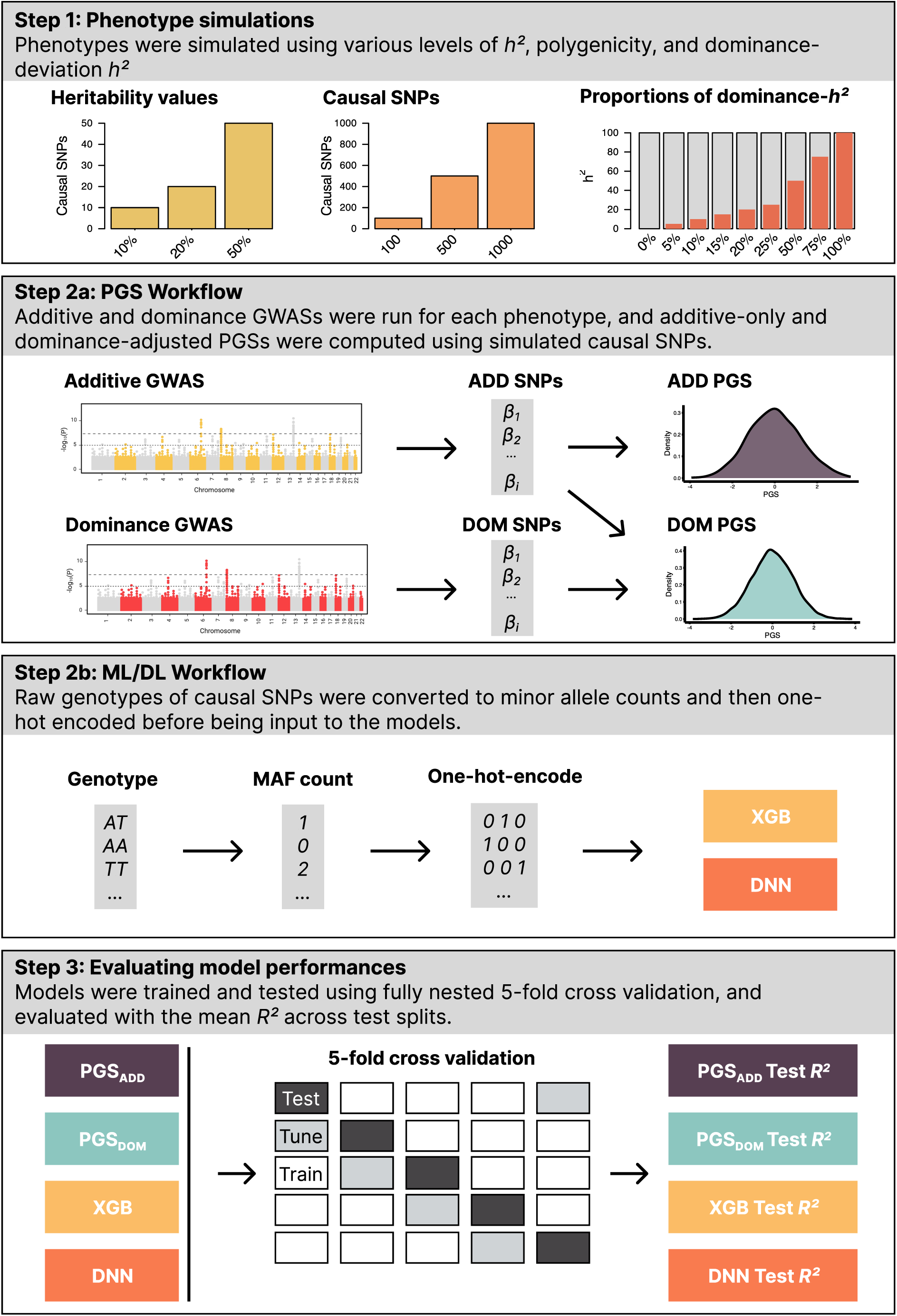
Graphic representation of simulation and prediction methods. Discovery (n = 100,000) and target (n = 50,000) genotype samples were simulated with HAPNEST using 1,329,052 autosomal SNPs. Synthetic phenotypes were simulated for these genotype samples, with varying proportions of *h*^2^ coming from dominance-SNPs. Additive and dominance GWASs were run per phenotype, and then the simulated causal SNPs were used for PGS construction and ML/DL model input. We tested the performance of four different models – an additive PGS, dominance-adjusted PGS, deep neural network, and XGBoost ensemble learner – using fully nested 5-fold cross validation.

### Impact of dominance at GWAS-like levels

Because dominance contributions to complex traits are generally modest (0.5% - 0.83%), we first assessed model performance under low-dominance conditions. We simulated phenotypes with 100 causal SNPs and varied the total SNP *h*^2^ (10%, 20%, 50%) and allocated 0 – 20% of SNP *h*^2^ to dominance-SNPs. In these conditions, additive PGS performance remained stable (Fig. 2a; Table S3), with a decline in mean *R*^2^ for the additive PGS of -0.007 when 20% of the total SNP *h*^2^ as attributable to dominance-SNPs. These results indicate that additive PGSs are robust to modest dominance contributions.

**Figure 2:**
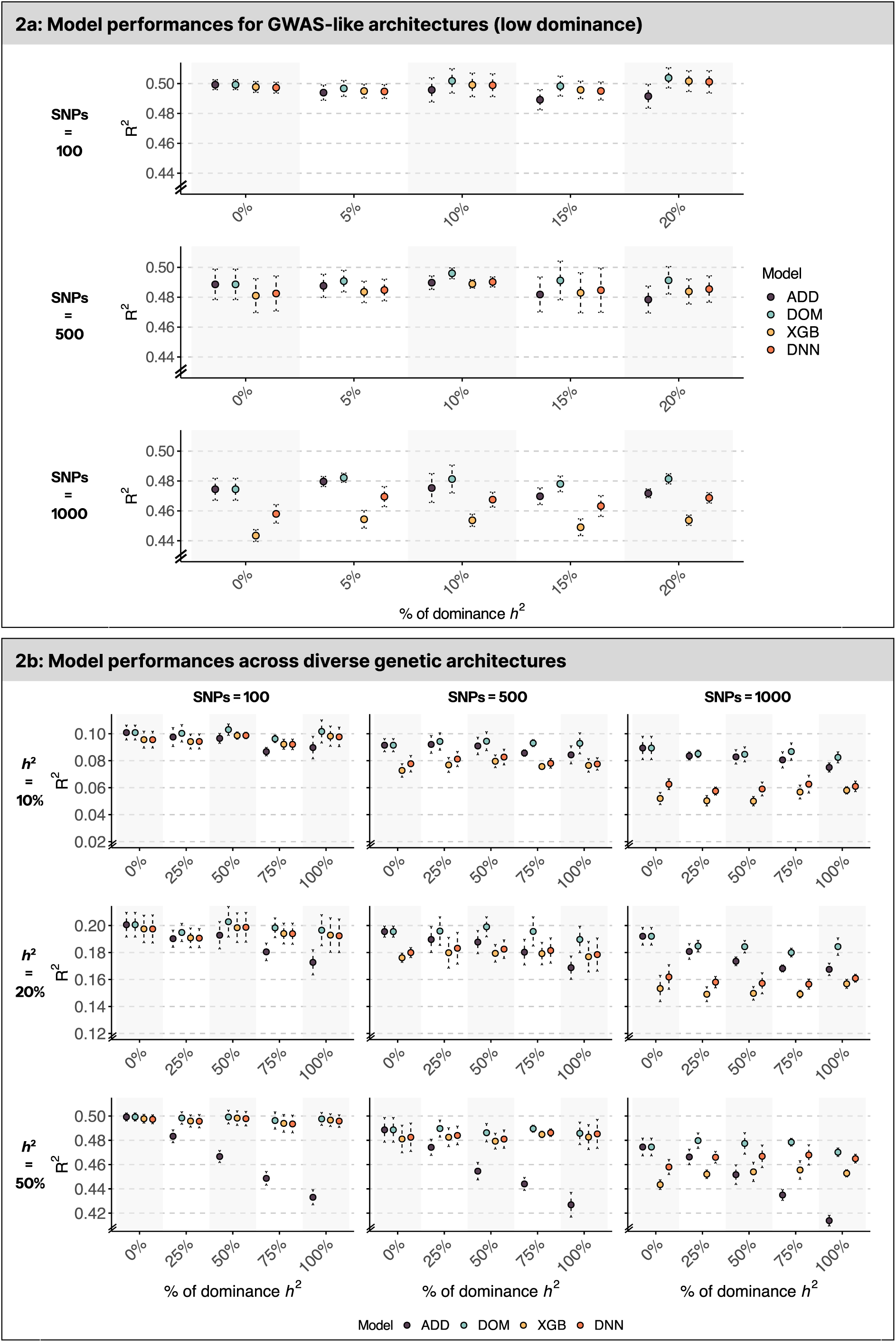
Model performances across different simulated genetic architectures. Panel **1a** shows the mean variance explained (*R*^2^) for phenotypes with a total SNP heritability of 50% in low dominance scenarios (0% - 20% of total SNP *h*^2^). Panel **1b** illustrates model performances across a wide range of simulated scenarios. Each point represents the mean variance-explained across 5 test splits (Y-axis; error bar = 1 SD). Columns represent differing amounts of polygenicity (i.e., number of causal SNPs) and rows represent different levels of total *h*^2^. Model codes are as follows: “ADD” = linear regression using an additive PRS; “DOM” = linear regression using a PRS adjusting for dominance deviations; “XGB” = XGBoost model using one-hot encoded raw genotype values; “DNN” = deep neural network using one-hot encoded raw genotype values. X-axis labels display percentages of the total SNP heritability caused by dominance-SNPs.

### Impact across diverse range of dominance, heritability and polygenicity

Having established that additive PGS performance is stable under GWAS-like, low-dominance conditions (Fig. 2a), we next examined a broader range of conditions by varying the proportion of dominance *h*^2^, total SNP *h*^2^, and polygenicity. Specifically, we evaluated predictive performance across three levels of total SNP heritability (*h*^2^ = 10%, 20%, 50%), each comprising five phenotypes with increasing proportions of *h*^2^ attributable to dominance-SNPs (dominance *h*^2^ = 0%, 25%, 50%, 75%, and 100%; Methods). As shown in Fig. 2b, the predictive performance of the additive PGS model declined as the proportion of dominance deviation-attributable *h*^2^ increased to high levels – particularly for traits with a high total SNP heritability. When total SNP *h*^2^ = 50%, performance dropped when ≥ ½ of the heritability arose from dominance-SNPs (median Δ*R*^2^ = -0.033). For *h*^2^ = 20%, a decline was observed when ≥ ¾ of *h*^2^ was due to dominance-SNPs (median Δ*R*^2^ = -0.02). For low-heritability phenotypes (*h*^2^ = 10%), decline in additive PGS performance was modest, with decreases occurring at higher levels of polygenicity and when high proportions of heritability were attributable to dominance-SNPs (median Δ*R*^2^ = -0.011). For all additive model results, see Table S3.

Compared to the additive PGS, the DNN and XGB models demonstrated improved performance in specific scenarios – particularly when heritability was high and polygenicity was low. However, as the number of causal SNPs increased and total heritability decreased, both ML/DL models exhibited substantial declines in predictive accuracy – even when the trait’s entire SNP *h*^2^ came from dominance-SNPs. For example, with total *h*^2^ = 10% and 1000 causal SNPs, the additive PGS outperformed both the DNN (Δ*R*^2^ = -0.014) and XGB (Δ*R*^2^ = -0.016) models. In contrast, the dominance-adjusted PGS consistently achieved performance close to the true simulated *h*^2^ across all architectures. Overall, model performance decreased as polygenicity increased, particularly for the ML/DL models, and the additive PGS decreased as the proportion of dominance *h*^2^ increased (for full results see Table S4).

### Impact of dominance deviation magnitude

To isolate how the size of the dominance deviation affects predictive performance, we held the total SNP *h*^2^, the additive SNP *h*^2^, and the dominance-SNP *h*^2^ constant at *h^2^_total_* = 50%, *h^2^_ADD_* = 37.5%, *h^2^_DOM_* = 12.5% (*h^2^_DOM_* is ¼ of *h^2^_total_*) and varied the strength of the dominance deviation (*k*). Values for *k* ranged from 0 (no dominance deviation) to -1.0 (total under-dominance; Fig. 3a). Figure 3b demonstrates that additive PGS performance declines as *k* gets further from zero (Table S5). Additionally, we see that total SNP heritability makes a difference on this decline, with the decreases in mean *R*^2^ of -0.018, -0.027, and -0.057 for total SNP *h*^2^ values of 10%, 20%, and 50% when *k* = -1.0, respectively. In contrast, the DOM PGS, XGB and DNN models were not affected by the magnitude of the dominance, which suggests that these methods are capable of accurately capturing the genetic risk of diseases with highly penetrant recessive variants (full results reported in Table S6).

**Figure 3:**
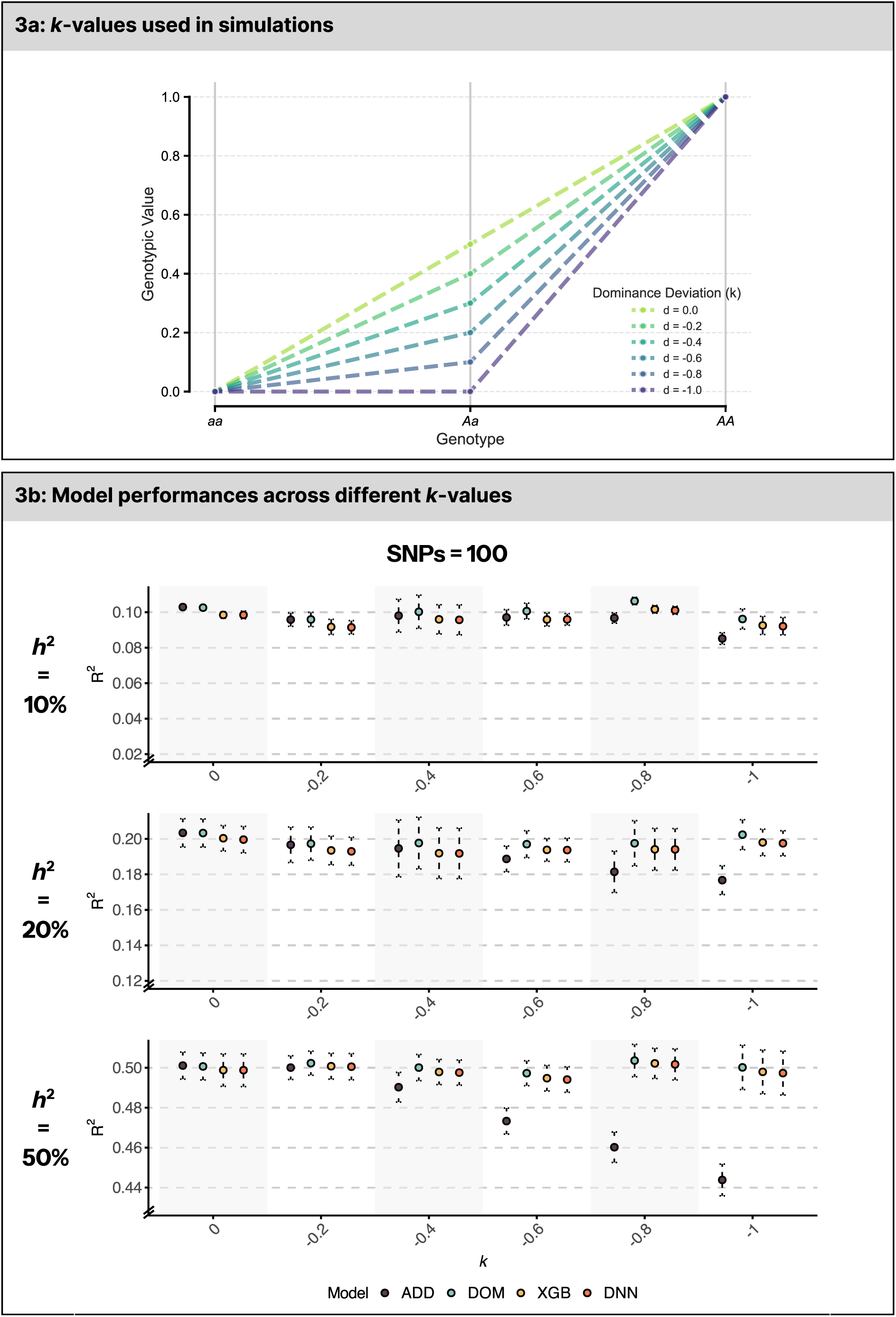
Model performances for different sized dominance deviations. Panel **3a** displays the dominance deviations of causal dominance-SNPs used for the simulated phenotypes in panel **3b**. Panel **3b** displays the mean variance-explained (Y-axis) values across 5 test splits of the four models as the magnitude of the dominance deviations of causal dominance-SNPs increase (from left to right). The X-axis displays size of dominance deviations for causal dominance-SNPs, ranging from *k* = 0 (no dominance deviation) to -1.0 (completely recessive). “ADD” = linear regression using an additive PRS; “DOM” = linear regression using a PRS adjusting for dominance deviations; “XGB” = XGBoost model using one-hot encoded raw genotype values; “DNN” = deep neural network using one-hot encoded raw genotype values.

### Performance in the UK Biobank

To assess model performance in real-world data, we applied the four predictive models to ten phenotypes in the UK Biobank (Fig. 4; Table S7). For each trait we conducted both additive and dominance GWASs using > 215,000 unrelated European individuals (discovery cohort; Table S7). Clumping and thresholding (C+T) was performed on the resulting summary statistics to select SNPs for computing PGSs and inputting into the ML/DL models (Fig. 4; Table S7; Methods). The prediction models were trained and tested in a non-overlapping target sample of 90,000 – 100,000 individuals (depending on phenotype; Table S7), using a fixed 70% training, 15% tuning, and 15% test split.

**Figure 4:**
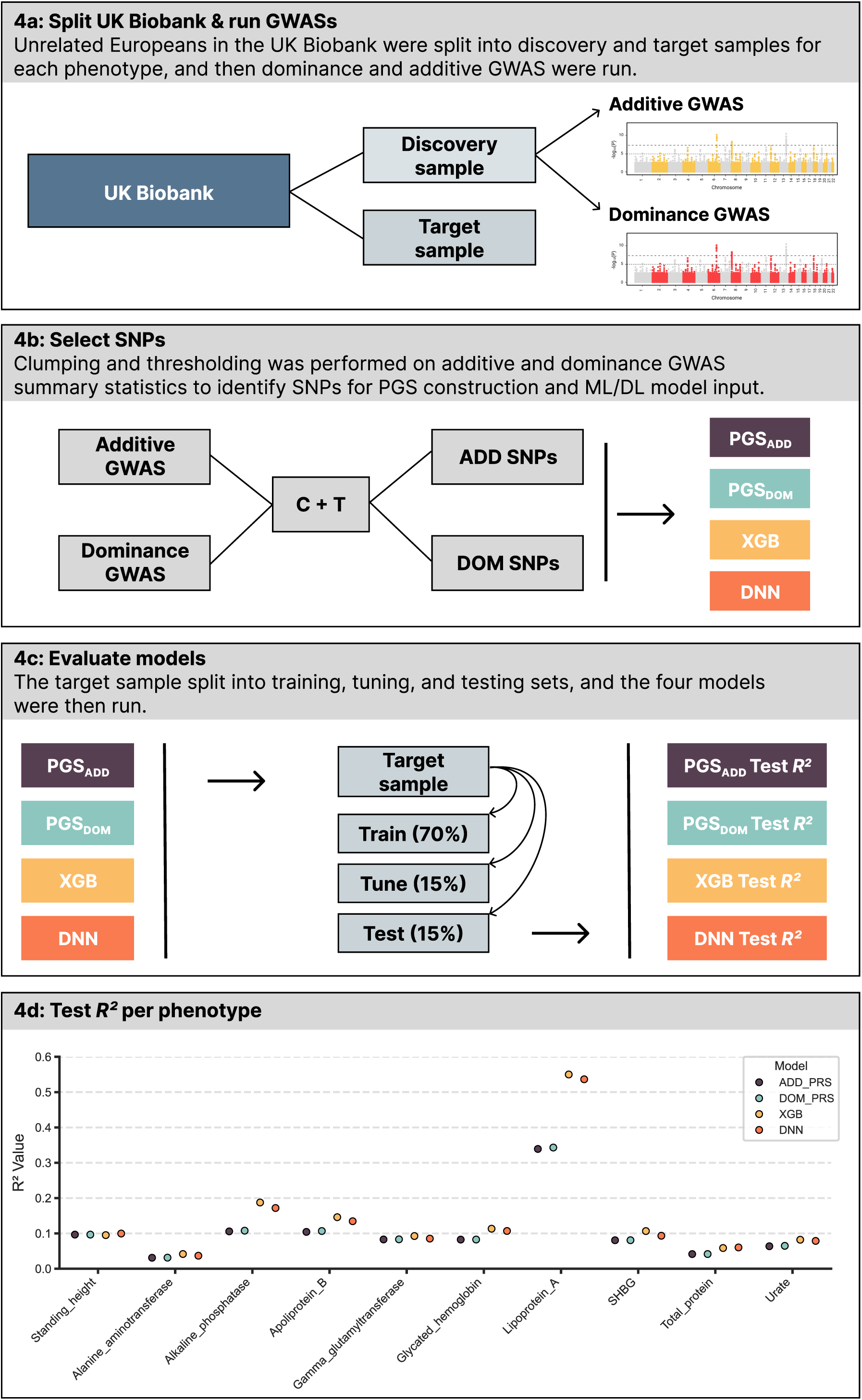
Methods and model performances in the UK Biobank. **(4a)** The UK Biobank was split into discovery and target samples for each of the ten phenotypes used (Table S7), and additive and dominance GWASs were performed (Methods). **(4b)** Clumping and thresholding (C+T) was used on both additive and dominance summary statistics to select SNPs to construct PGSs and input into ML/DL models, which was performed using the same methods as the simulation models. **(4c)** The target samples were split using a 70% train, 15% tune, 15% test split. **(4d)** The variance-explained values on the held-out test set are shown for the four predictive models. “ADD” = linear regression using an additive PRS; “DOM” = linear regression using a PRS adjusting for dominance deviations; “XGB” = XGBoost; “DNN” = deep neural network.

Of the ten phenotypes analysed, nine were selected based on prior evidence of dominance effects at genome-wide significant loci, while standing height served as a negative control with no loci with known dominance deviations^16^. As expected, all four models performed similarly for standing height (Fig. 4d). In contrast, for the nine phenotypes with documented dominance deviations, the DNN and XGB models outperformed the additive and dominance-adjusted PGS models. The largest gains in *R*^2^ were observed for lipoprotein(a) and alkaline phosphatase, where XGBoost improved the variance explained by 0.211 and 0.081, respectively, relative to the additive PGS. Additional improvements were seen for apoliprotein B (Δ*R*^2^ = 0.0416), glycated haemoglobin (Δ*R*^2^ = 0.0308), and SHBG (Δ*R*^2^ = 0.0259), with comparable performance from the DNN model. Notably, the dominance-adjusted PGS did not match the performance of either ML/DL approach, suggesting that simple dominance extensions to additive scores may be insufficient to fully capture non-additive effects in real-world data. This may be due to several reasons, including the ratio of additive to dominance-SNPs when selecting SNPs for PGS construction and ML/DL model input, or possibly that the ML/DL methods capture non-additive effects not explicitly modelled in the dominance-adjusted PGS. For full model results in UK Biobank phenotypes see Table S7.

To summarize our findings, we created a decision framework outlining when additive PGS performance may suffer due to dominance deviations, and when the use of ML/DL approaches may be warranted (Fig. 5). This flowchart synthesizes results from both the simulations and UK Biobank analyses, providing practical guidance for model selection depending on the genetic architecture of the trait in question.

**Figure 5:**
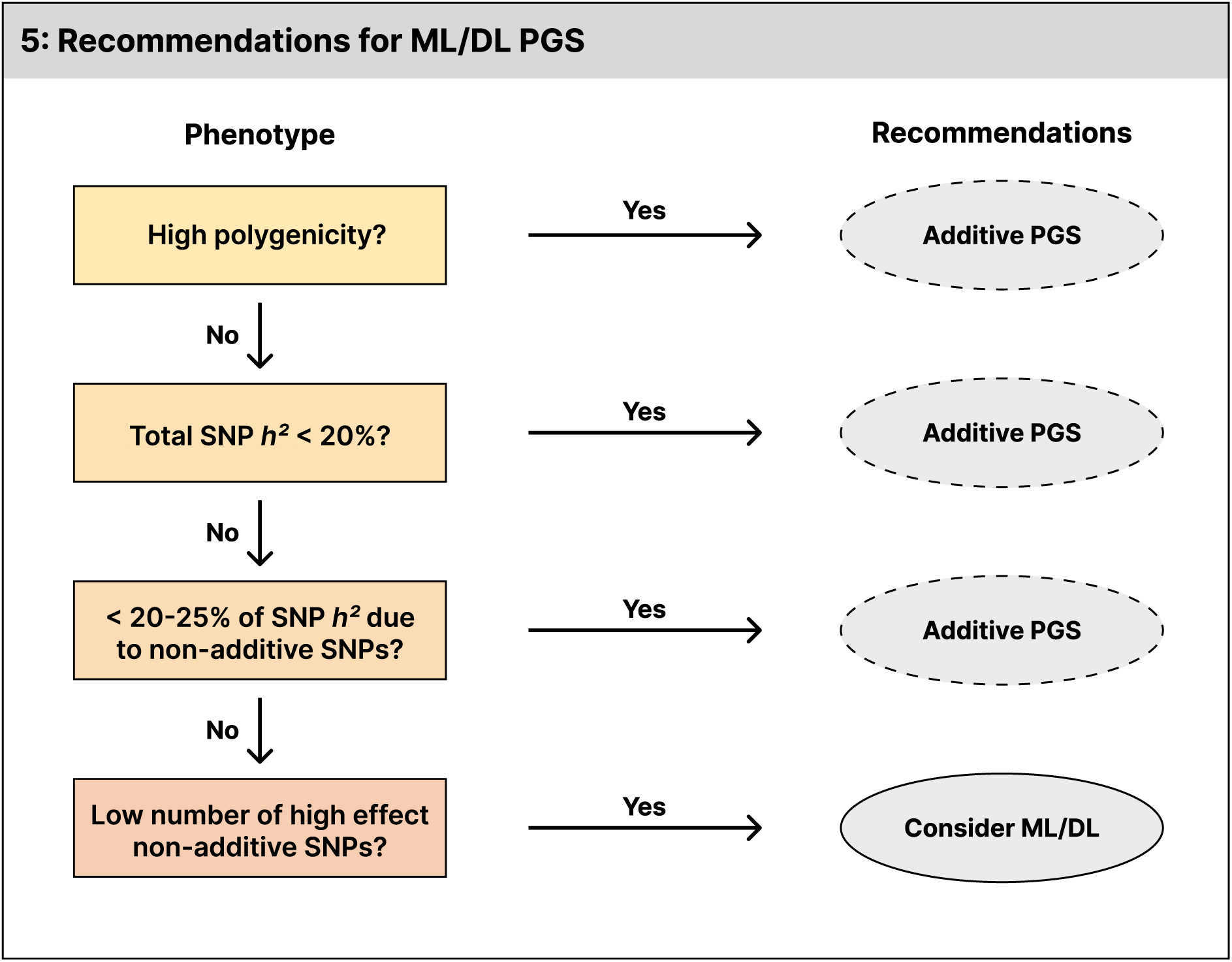
Summary of recommendations for using ML/DL with varying genetic architecture. Flow chart illustrating recommended approaches for when to consider using ML/DL in the place of additive PGS depending on the genetic architecture of the phenotype. Under most conditions, additive PGS models are generally sufficient and robust to small amounts of dominance. The use of ML/DL models should be primarily considered when the phenotype is question has a high total SNP heritability, low levels of polygenicity, a high portion of SNP heritability attributable to dominance-SNPs, and/or dominance-SNPs with very large effects.

## Discussion

Machine learning and deep learning methods for genetic prediction have been suggested to better model non-additive genetic effects that are potentially overlooked by standard PGS approaches. In this study, we systematically evaluated how one form of non-additive genetic effects – specifically, recessive dominance deviations – impact the performance of additive PGSs, and whether ML/DL models or dominance-adjusted PGSs can offer alternatives in these conditions. Using both simulated phenotypes and real-world data from the UK Biobank, we compared the predictive performance of four models: an additive-only PGS, a dominance-adjusted PGS, a deep neural network, and an XGBoost ensemble learner.

Our findings demonstrate that dominance deviations can impair the accuracy of additive PGSs, particularly for traits with high SNP *h*^2^, a substantial proportion (≥ 25%) of *h*^2^ due to dominance-SNPs, low levels of polygenicity, and strong dominance deviations. The ML/DL models outperformed additive PGS only in conditions with low polygenicity and high SNP heritability; under more polygenic and lower-heritability scenarios the additive PGS remained more accurate. Across all simulated architectures, the dominance-adjusted PGS performed close to the theoretical maximum *R*^2^. In contrast, when applied to real traits in the UK Biobank, ML/DL models outperformed both additive and dominance-adjusted PGSs for phenotypes with known dominance effects, while all models performed consistently for standing height – a control trait with no known dominance deviations.

While our simulations were designed to mimic dominance deviations of equal effect across the genome, they do not exactly mirror the architecture of the dominance observed in the real-world data from the UK Biobank – which tend to have a smaller number of large effect loci. Instead, we used a simulation design that can be interpreted as a conservative or “worst case” scenario – that many small and difficult to detect dominance deviations are sufficient to degrade additive PGS performance. Our results suggest that in most cases they are not: additive PGS remained robust across most simulation settings, with performance only declining under conditions of high SNP *h*^2^, low polygenicity and substantial proportions of heritability attributable to dominance-SNPs. For reference, we observe minimal to no decline in additive PGS performance when ¼ of the *h*^2^ was attributable to dominance-SNPs, which is a sizeable proportion considering the average ratio of additive to dominance heritability is noted to be between 0.5% and 0.83%^16–18^. Our findings indicate that widespread, modest dominance effects do not necessitate the use of more complex modelling in most cases.

However, the model results from the UK Biobank provide insight into the specific contexts where ML/DL models may offer superior predictive accuracy. Across the nine traits with known dominance effects, both the deep neural network and XGBoost models outperformed the additive and dominance-adjusted PGSs – despite all models using the same sets of SNPs. These improvements were most pronounced for lipoprotein(a) and alkaline phosphatase, both of which are characterized by medium to high SNP heritability and very few strong effect GWAS loci. In contrast, all models performed similarly for standing height, a highly polygenic trait with no known dominance effects. Together with our simulation results, these findings suggest the ML/DL methods may be beneficial for a narrow subset of traits: those with low polygenicity, high heritability, and strong dominance deviations (e.g., biomarker and/or eQTL traits). For most complex traits, particularly those with diffuse genetic architectures or modest SNP heritability, additive PGS methods remain the most robust and practical choice.

The discrepancy between the simulation and UK Biobank results – namely, that the dominance- adjusted PGS did not perform comparably to ML/DL in the UK Biobank – may reflect differences in SNP selection and genetic architecture. In simulations, we tested models using the exact causal variants to assess how models performed in the most ideal scenario. However, SNPs for the UK Biobank models were selected using C+T on both the additive and dominance GWAS summary statistics. Even for lipoprotein(a) – the phenotype with the strongest dominance effects - only 20 lead dominance-SNPs were identified, in comparison to 770 lead additive SNPs. This disparity highlights the challenge of accurately capturing dominance signal using GWAS. Consequently, the dominance-adjusted PGS may underperform not because of an inadequate modelling framework but because of either inaccurate estimation of weights of dominance effects in comparison to their additive counterparts, or the need for more extensive tuning to identify the optimal parameters for determining when a SNP has a dominance deviation. It is possible that (i) further refinement of p-value and LD cutoff thresholds for C+T, or (ii) larger sample sizes could improve predictive accuracy of the dominance-adjusted PGS. Additionally, it is possible that the ML/DL models are capturing non-additive effects not explicitly modelled by the dominance-adjusted PGS. Overall, our results highlight the potential value of a hybrid approach of combining additive PGS with raw genotypes for known dominance loci within an ML/DL model. This type of strategy may improve predictive performance without the extensive training and tuning needed for models built solely upon raw genotype values.

In summary, we systematically evaluated the impact of dominance deviations on polygenic prediction by comparing additive PGS, dominance-adjusted PGS, a neural network and an XGBoost ensemble learner in both simulated and real-world data from the UK Biobank. Our findings demonstrate that dominance effects can reduce the accuracy of additive PGSs, particularly for traits with high SNP heritability, low polygenicity, and large-effect dominance deviations. In such cases, ML/DL models and dominance-adjusted PGSs can offer improved predictive performance. However, for the vast majority of traits – especially as polygenicity rises and heritability decreases – additive PGSs remain more robust and practical. While ML/DL methods may be useful for a subset of phenotypes characterized by sparse, strong-effect dominance architecture, our findings suggest that widespread, modest dominance deviations do not, on their own, justify the routine use of complex ML/DL modelling for genetic-based predictions.

## Methods

### Genotype simulations

We simulated two synthetic genotype samples of European descent using HAPNEST: a discovery sample (*n* = 100,000) and a target sample (*n* = 50,000). HAPNEST generates genotypes by resampling reference genomes with a stochastic model that estimates processes like recombination and mutation^31^. The 1000 Genomes Project (1000G) European reference panel was used for the simulations, and both samples contained genotypes with 1,329,052 SNPs across the 22 autosomal chromosomes. No X or Y chromosomes were simulated – all individuals in the samples were sex ambiguous.

### Phenotype simulations

We simulated quantitative phenotypes on synthetic HAPNEST genotypes under controlled additive and dominance architectures. First, to reflect typical GWAS-like levels of dominance, we set total SNP-heritability = 50% with 100, 500, and 1000 causal SNPs, dominance *h*^2^ ranging from 0 – 20% (as a fraction of total *h^2^_SNP_*). Additive causal SNPs were given effects exactly at the midpoint between the homozygotes (*aa* = 0, *Aa* = 0.5, *AA* = 1; dominance deviation [*k*] = 0), and non-additive SNPs were given heterozygote effects 0.5 times less than expected (*aa* = 0, *Aa* = 0.25, *AA* = 1; *k* to -0.5). We then ran a full grid of simulations with total *h^2^_SNP_* values of 10%, 20%, and 50%, with 100, 500, and 1000 causal SNPs, dominance *h*^2^ ranging from 0 – 100% of total *h^2^_SNP_* and using the same *k* values as above for non-additive and additive causal SNPs. Finally, to isolate the effects of dominance deviation magnitude, we used *h^2^_SNP_* values of 10%, 20%, and 50%, fixed causal SNPs to 100 and the dominance *h*^2^ to 25% (of total *h^2^_SNP_*), and varied values of *k* from 0 to -1.0. For each configuration, additive and non-additive SNP effects were summed and then scaled to ensure their true *R*^2^ values would meet the target for the given phenotype. Phenotype simulations were validated using linear regression, to ensure that the *R*^2^ for additive, non-additive, and combined effects all matched the expected values. To account for minor allele frequency (MAF), SNPs were separated into deciles and then randomly selected using the sample() function in R to ensure an even distribution across MAF bins. The phenotype simulations successfully achieved the desired partitions of additive, non-additive, and combined SNP *h*^2^ , as confirmed with linear regressions of the true genetic effects for each phenotype (Table S1 and S2).

### Simulation GWASs

For each of the simulated phenotypes, two GWASs were run: one assuming only additive SNP effects, and a second modelling dominance deviations as well. For both GWASs, the first 10 principal components (PCs) were corrected for; given that the simulated individuals in our sample were “sex ambiguous” and have no age these were not included as covariates. For the additive GWAS, synthetic phenotypes were simply regressed on the SNP risk allele dosage and first 10 PCs, as typically done. For the dominance deviation GWAS, a dominance deviation component was added into the linear regression for each SNP, with homozygotes coded as 0 and heterozygotes coded as 1. This allows the effect size and significance of the dominance deviation to be assessed and was done using the “—glm genotypic” flag in PLINK. All GWASs were run with PLINK v2.00a6^32^.

### UK Biobank data

Data for real phenotype analyses in this study were obtained from the UK Biobank, a prospective population-based cohort, under application number 16406. Participation in the UK Biobank is voluntary, and each participant provided written informed consent. Ethics approval for the UK Biobank was granted by the National Research Ethics Service Committee North West-Haydock (reference 11/NW/0382). The resource provides extensive individual-level longitudinal data that collects lifestyle, physical, biological, genetic, imaging, and electronic health record data. This study analyzed individuals of European descent, with a mean age of 56.72 (SD = 8.03) years, with 53.8% females across the phenotypes used (see Table S7 for demographic information per phenotype). Subjects were randomly assigned per phenotype into either the discovery or replication cohort.

Genetic data from the UK Biobank used in this study were genotyped with the Applied Biosystems UK Biobank Axiom Array, covering 825,927 SNPs. Quality control, haplotype phasing, and imputation were performed by the UK Biobank, and are described elsewhere^33^.

### UK Biobank GWASs

Phenotypes from the UKB were selected based on prior evidence for at least one causal SNP exhibiting a dominance deviation, as seen in Palmer et al^16^. In addition to these phenotypes, standing height was included as a control phenotype. We extracted the phenotypes using the following field IDs: glycated haemoglobin (30750), Urate (30880), Alkaline phosphatase (30610), Alanine aminotransferase (30620), Gamma glutamyltransferase (30730), SHBG (30830),

Lipoprotein A (30790), Total protein (30860), apoliprotein B (30640), Standing height (50). We analysed EUR ancestry individuals after empirically assigning individuals to ancestral continental populations using the 1000 Genomes reference panel based on Mahalanobis distance of the first 20 Principal components (PCs). PCs were created using FlashPCA2. Prior to running GWASs, all UKB phenotypes were transformed using inverse-rank normalization, as done elsewhere^16^. As done for the simulations, two separate GWASs were conducted per phenotype: one with a purely additive model (“—glm” in PLINK 2.0) and one that models dominance deviations (“—glm genotypic”). We included sex, age, array, batch, and the first 10 within EUR ancestry PCs as covariates after standardization (--covar-variance-standardize). We analysed hard-called imputed variants with an INFO score >0.9. This resulted in between 200 and 300k individuals and 16.7 variants (Table S7).

### SNP selection for PGS construction and ML/DL model input

For all simulated phenotypes, SNPs selected for PGS construction and input into ML/DL models was based on their true effect status (i.e., causal additive SNP or causal dominance-SNP). For UK Biobank analyses, SNPs were selected using C + T with the following parameters: *--*clump-p1 = 0.000001, --clump-p2 = 0.0001, --clump-r2 = 0.2, and--clump-kb = 250. Clumping and thresholding was performed using PLINK v1.90b7.

### PGS models

Two different PGS models were tested for each of the simulated phenotypes: a linear regression using an additive PGS, and a linear regression using a PGS incorporating additive and dominance deviation effects. The additive-only PGS was computed using the standard formula:

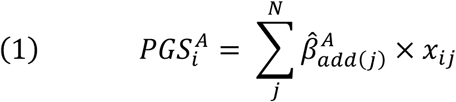

where *i* represents the individuals, *N* represents the numbers of SNPs, *j* represents each SNP, 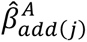 is the estimated effect for SNP *j* from the discovery GWAS assuming additive genetic risk, and *x*_*i*j_ refers to the number of risk increasing alleles for a given SNP. The adjusted dominance deviation PGS was computed by accounting for the dominance deviation using the following formula:

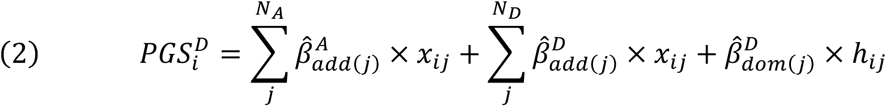

where *N*_*A*_ is the number of causal SNPs with only additive effect, *N*_*D*_ the number of causal SNPs with a dominance effect, 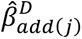 and 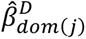 are the estimated additive and dominance effect for SNP *j* from the discovery GWAS incorporating dominance deviation, and *h*_*i*j_ = 1 equals one if *x*_*i*j_ = 1, and *h*_*i*j_= 0 otherwise. When calculating the dominance deviation-adjusted PGS, the risk score for non-additive causal SNPs were computed using formula 2, whereas as additive causal SNPs were computed using formula 1. Per-SNP scoring followed the simulation’s ground truth architecture: SNPs labelled additive were scored formula (1), and SNPs with a dominance deviation were scored with formula (2).

### Deep neural networks

Neural networks are powerful tools for modelling complex, non-linear relationships in data, and have successfully improved prediction for biological questions such as antimicrobial resistance and trait-associated gene expression^34,35^. A feedforward neural network is a supervised machine learning model that maps input features (in this study, SNP genotypes) to an outcome (e.g., a phenotype) through multiple layers of weighted connections and non-linear transformations, allowing to capture complex relationships in data. The neural networks for this research were implemented in PyTorch (v2.0.1) to predict the synthetic genetic phenotypes using one-hot encoded SNP risk allele dosages as input^36^. The models’ architecture consisted of 2 fully connected hidden layers with 512, and 128 neurons, respectively. The exponential linear unit (ELU) activation function was used in all layers to ensure smooth gradients and prevent dying neurons^37^. Dropout was applied to the first hidden layer to prevent overfitting and help with generalization. The AdamW algorithm was used as an optimizer and mean squared error (MSE) was used as the loss function^38^. DNN models were trained on the Dutch Snellius high performance computing (HPC) cluster, using a single NVIDIA A100 GPU.

### Extreme gradient boosting

XGBoost is a supervising machine learning algorithm that builds an ensemble of decision trees in a sequential “boosting” framework, where each tree corrects the errors of previous ones. Similar to neural networks, it is capable of modelling non-linear relationships, with previous research demonstrating it can learn complex genetic effects and allele interactions in genetic-based phenotypes^28,39^. The XGBoost regression models used here were run using the Python xgboost package (v2.1.3) and took the one-hot encoded SNP risk allele dosage for the known causal SNPs as input^39^. Like the DNNs, the MSE loss function was used, and a single NVIDIA A100 GPU was used to accelerate computation.

### Hyperparameter tuning & model assessment

Machine and deep learning models can require extensive tuning of hyper parameters to maximize performance. To do this, we used fully nested 5-fold cross validation, where the data is split into 5 partitions which are cycled through so each is used once as the hold out test set. The mean *R*^2^ is computed across test sets to assess model performance. The Optuna package (v4.1.0) was used in Python to search for the optimal hyper parameters: learning rate, dropout probability, and L2 regularization values for the DNNs and learning rate, minimum child weight, subsample ratio of columns, and L1 and L2 regularization values for XGBoost^40^.

## Data Availability

Code and data for simulated phenotypes and analysis are available on GitHub [https://github.com/nybell/non-add-paper]. Code for all UK Biobank analyses can be found on the same GitHub, and a description of the data can be found on the UK Biobank website, while the data itself can be accessed on the UK Biobank Research Analysis Platform for approved researchers [https://www.ukbiobank.ac.uk/use-our-data/research-analysis-platform/].

## Acknowledgments

This project was funded by European Union’s Horizon 2020 research and innovation programme under grant agreement No 964874. DP is supported by the Netherlands Organization for Scientific Research - Gravitation project ‘BRAINSCAPES: A Roadmap from Neurogenetics to Neurobiology’ (024.004.012), and by the European Research Council advanced grant ‘From GWAS to Function’ (ERC-2018-ADG 834057). NYB, DP, CdL and DP have no conflicts of interest to declare.

